# Antibody-mediated neutralization of authentic SARS-CoV-2 B.1.617 variants harboring L452R and T478K/E484Q

**DOI:** 10.1101/2021.08.09.21261704

**Authors:** Alexander Wilhelm, Tuna Toptan, Christiane Pallas, Timo Wolf, Udo Goetsch, Rene Gottschalk, Maria JGT Vehreschild, Sandra Ciesek, Marek Widera

## Abstract

The capacity of convalescent and vaccine-elicited sera and monoclonal antibodies (mAb) to neutralize SARS-CoV-2 variants is currently of high relevance to assess the protection against infections.

We performed a cell culture-based neutralization assay focusing on authentic SARS-CoV-2 variants B.1.617.1 (Kappa), B.1.617.2 (Delta), B.1.427/B.1.429 (Epsilon), all harboring the spike substitution L452R.

We found that authentic SARS-CoV-2 variants harboring L452R had reduced susceptibility to convalescent and vaccine-elicited sera and mAbs. Compared to B.1, Kappa and Delta showed a reduced neutralization by convalescent sera by a factor of 8.00 and 5.33, respectively, which constitutes a 2-fold greater reduction when compared to Epsilon. BNT2b2 and mRNA1273 vaccine-elicited sera were less effective against Kappa, Delta, and Epsilon compared to B.1. No difference was observed between Kappa and Delta towards vaccine-elicited sera, whereas convalescent sera were 1.5-fold less effective against Delta, respectively. Both B.1.617 variants Kappa (+E484Q) and Delta (+T478K) were less susceptible to either casirivimab or imdevimab.

In conclusion, in contrast to the parallel circulating Kappa variant, the neutralization efficiency of convalescent and vaccine-elicited sera against Delta was moderately reduced. Delta was resistant to imdevimab, which however, might be circumvented by a combination therapy with casirivimab together.

## 1. Introduction

In RNA viruses, such as the severe acute respiratory syndrome coronavirus 2 (SARS-CoV-2), mutations occur during their replication by substitution, insertion or deletion of nucleotides in the viral genome [1]. In most cases, silent mutations have no impact on protein structure and function. However, certain amino acid changes in the region coding for the spike protein (S), may not only affect the protein function but also alter its immunogenic capacity [2]. SARS-CoV-2 S binds human ACE2 receptor and is subsequently cleaved by TMPRSS2 transmembrane protease to enter the host cell to initiate replication.

S is the major immunogenic compound of mRNA and vector-based vaccines [3]. Furthermore, the formation of antibodies against the spike protein neutralizes the SARS-CoV-2 S and protects against infection. In combination with cellular responses vaccination protects against severe COVID-19 disease [4, 5]. Changes in the structure, however, could reduce the effectiveness of vaccines as the current generation of mRNA and vector-based vaccines were developed against the spike protein of the Wuhan-Hu-1 isolate. Similarly, most of the commercially available monoclonal antibodies used for prevention and therapy were released in 2020. Due to emerging SARS-CoV-2 variants, there is an eminent interest in evaluating mutations in S for potential immune escape.

Variant Alpha (B.1.1.7) S protein binding to the human ACE2 receptor with increased affinity is most probably responsible for the higher transmission rate [6]. Alpha rapidly became the predominant variant in UK (https://cov-lineages.org/global_report_B.1.1.7.html) and spread globally as a result of international travel, which is the major driver of the introduction and spread of SARS-CoV-2 variants [7, 8]. Currently, Alpha is displaced by the Delta variant (B.1.617.2), which was first identified in India in late 2020. Delta has since dominated over other sublineages including B.1.617.1 (Kappa) [9], which was found earlier in India in 2020. Potential reason for globally attained dominance of delta is believed to be a higher transmissibility and immune evasion [10, 11]. While Alpha, Beta and Gamma all harbor the N501Y substitution in S, Delta and many variants of interest (VOI) such as Epsilon or Zeta gained other mutations e.g. the L452R[12]. The Epsilon lineages B.1.427 and B.1.429 originated in California and differ only in Orf1a and Orf1b, but carry identical mutations in S. Increased infectivity observed *in vitro* is in line with the progressive spread of Epsilon into other countries [13]. It has been shown that SARS-CoV-2 variants carrying E484K have limited susceptibility to convalescent and vaccine-elicited sera as well as monoclonal antibodies *in vitro* [14]. Moreover, E484K located within the S-ACE2 interface contributes to increased affinity to ACE2 resulting in enhanced virulence of variant Beta and Gamma [15-17]. In Kappa, but not Delta, E484 is substituted with a Glutamine (Q) and might confer immune escape similar to E484K [18]. In Delta, in close proximity to E484, a threonine is replaced by a positively charged lysine leading to T478K [19]. Little is known so far about the clinical relevance of the newly emerging T478K substitution. Whether T478K or E484Q, respectively, contribute to immune evasion similar to the E484K mutation is of great therapeutic importance as well as for evaluating the efficacy of currently approved vaccines. In this study we evaluated the antibody-mediated neutralization of authentic SARS-CoV-2 variants Kappa and Delta in comparison to FFM1 (B), FFM7 (B.1), Alpha and Epsilon strains against vaccine-elicited serum samples after immunization with BNT162b2 and mRNA1273, convalescent sera as well as mAbs bamlanivimab and casirivimab/imdevimab.

## 2. Materials and Methods

### 2.1 Cell culture and virus propagation

Caco2 and A549-AT cells [20] were cultured in Minimum Essential Medium (MEM) supplemented with 10% fetal calf serum (FCS), 4 mM L-glutamine, 100 IU/ml of penicillin and 100 μg/ml of streptomycin at 37°C and 5% CO2. The Caco2 cells were obtained from DSMZ (Braunschweig, Germany, no: ACC 169) and selected for high permissiveness to SARS-CoV-2 infection by serial dilution and passaging. SARS-CoV-2 isolates were grown using Caco2 cells as described previously [7]. Cell free cell culture supernatant containing infectious virus was harvested after complete cytopathic effect (CPE) and aliquots were stored at -80°C. Titers were determined by median tissue culture infective dose (TCID50) method as described by Spearman [21] and Kaerber [22] using Caco2 cells. GenBank accession numbers for the strains used in this study are as follows: B (FFM1/2020), MT358638, [23]; B (FFM5/2020), MT358641, [23]; B.1 (FFM7/2020), MT358643, [23]; B.1.1.7 (FFM-UK7931/2021), MZ427280, [14]; B.1.427 (FFM-CAL6541/2021), MZ317895; B.1.429 (FFM-CALsprt/2021), MZ317896.2; B.1.617.1 (FFM-IND5881/2021), MZ315140; B.1.617.2 (FFM-IND8424/2021), MZ315141. All virus-containing samples were inactivated according to standardized methods as described previously [24].

### 2.2 SARS-CoV-2 Neutralization Assay

SARS-CoV-2 antibody concentrations of this cohort were tested previously [14] with the SARS-CoV-2 IgG II Quant assay (Abbott Diagnostics) performed on the Alinity I with an analytical measurement range from 2.98–5680 binding antibody units per mL (BAU/mL).

Serum samples from convalescent and mRNA-1273 or BNT162b2 vaccinated individuals were serially diluted (1:2) and incubated with 4000 TCID50/mL of the indicated SARS-CoV-2 variant. Infected cells were monitored for cytopathic effect (CPE) formation after 72-hour inoculation. Values represent reciprocal dilutions of SARS-CoV-2 microneutralization titers resulting in 50% virus neutralization (NT50). The indicated monoclonal antibody solutions bamlanivimab, imdevimab and casirivimab were used in physiological concentrations according to the manufacturer’s instructions. For the evaluation of the neutralization capacity of mAbs, physiological concentrations according to the manufacturer’s instructions of monoclonal antibody solutions bamlanivimab, imdevimab, casirivimab and casirivimab/imdevimab (1:1) were serially diluted (1:2) and incubated with 4000 TCID50/mL of the indicated SARS-CoV-2 variant. CPE was evaluated microscopically after 72-hour inoculation.

### 2.3 Statistical analysis

Data analysis was performed in Microsoft Excel and GraphPad Prism 8 (GraphPad Software, USA). Statistical significance compared to untreated control was determined using unpaired Student’s t test on non-log-transformed data. Asterisks indicated p values as *p<0.05, **p≤0.01, and ***p≤0.005.

## 3. Results

### 3.1. Limited neutralization of SARS-COV-2 variants by convalescent and vaccine-elicited sera

In order to assess the susceptibility of SARS-CoV-2 to neutralizing antibodies, we tested convalescent and vaccine-elicited sera samples after immunization with BNT162b2 and mRNA1273. We found that the tested sera were less effective against authentic SARS-CoV-2 variants harboring L452R.

Convalescent sera showed reduced neutralization of Kappa and Delta by a factor of 8.00 and 5.33, respectively, relative to B.1 (**Fig. 1A**). For the Epsilon lineages B.1.427 and B.1.429 harboring D614G and L452R, the reduction in neutralization efficiency by convalescent sera of both variants was less severe with only 2.05 and 2.96, respectively. The serum from a immunocompromised patient which was long-term PCR positive for SARS-CoV-2 neutralized Kappa less efficiently compared to Delta and the Epsilon lineages.

**Figure 1.**
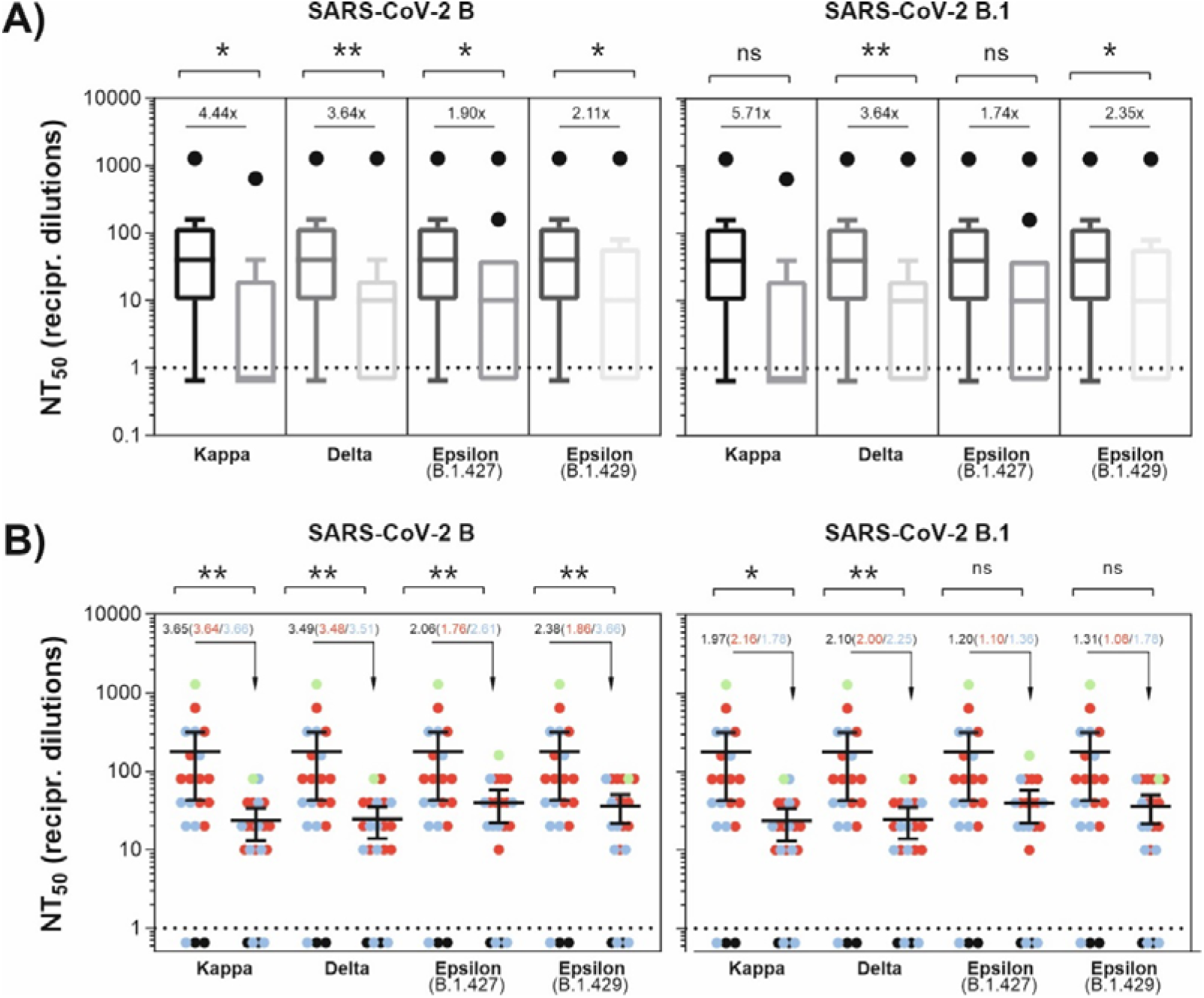
Antibody-mediated neutralization efficacy against SARS-CoV-2 variants. **(A)** A549-AT cells were incubated with serially diluted (1:2) sera from convalescent plasma with the indicated SARS-CoV-2 variant. Dots indicate convalescent sera (grey), negative sera (black) and a serum from an immunocompromised individual with a lasting SARS-CoV-2 infection (green). **(B)** A549-AT cells were incubated with serially diluted (1:2) sera from vaccinated individuals together with the indicated SARS-CoV-2 variant. Dots indicate individual sera from BNT162b2 (blue), mRNA1273 (red) vaccinated individuals and SARS-CoV-2 negative sera (black). Additionally, a serum from a convalescent and BNT162b2-vaccinated individual (green) is shown. Statistical significance compared to FFM1 (B) and FFM7 (B.1) was calculated by two-tailed, paired student’s t-tests. Green dots (A+B) were excluded from significance testing. Mean values are depicted from two replicates. Asterisks indicate p-values as * (p < 0.05), ** (p ≤ 0.01), and *** (p ≤ 0.001).

In presence of L452R, limited neutralization efficacy to BNT2b2 or mRNA1273 vaccine-elicited sera was detected (Fig. 1B). Both mRNA vaccine elicited sera neutralized Kappa and Delta 2-fold weaker than B.1, whereas B.1.427 and B.1.429 were less sensitive by a factor of 1.20 and 1.31, respectively (**Fig. 1B**). For both sera, the neutralization capacity against the B.1617 strains were 2-fold reduced compared to B.1.427 and B.1.429 (**Fig. 1A and B**). Kappa and Delta were equally sensitive towards vaccine-elicited sera (**Fig. 1B**). However, using convalescent sera, neutralizing titers obtained for Kappa versus Delta were 1.2-fold (cf. B) or 1.6-fold (cf. B.1) reduced, respectively (**Fig. 1A**).

### 3.2. Relative resistance of SARS-CoV-2 variants to monoclonal antibodies

Next, we sought to find out whether the efficacy of mAbs bamlanivimab, casirivimab, imdevimab, and the combination of casirivimab/imdevimab were reduced towards authentic SARS-CoV-2 variants harboring L452R and other substitutions in S. As evaluated in A549-AT cells [20], authentic SARS-CoV-2 variants harboring L452R were resistant against bamlanivimab (**Fig. 2**). In agreement with previously published data [14], all applied mAbs could efficiently neutralize variants B, B.1, and Alpha (**Fig. 2**). Kappa additionally harboring E484Q revealed strongly reduced susceptibility against casirivimab, which was approx. 5.5-fold lower compared to D614G alone (cf. B.1). Neutralization of Kappa by imdevimab or a combination therapy with casirivimab/imdevimab together was slightly reduced relative to non E484Q harboring variants. In contrast to Kappa, testing the Delta variant revealed a substantial resistance against imdevimab while casirivimab-mediated neutralization was still effective (**Fig. 2**).

**Figure 2.**
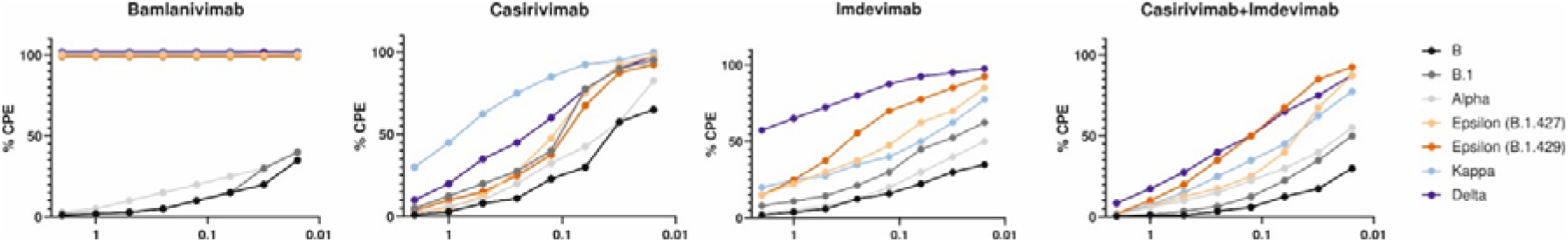
Neutralization efficacy of monoclonal antibodies against SARS-CoV-2 variants. Serially diluted (1:2) monoclonal antibodies (mAbs) were incubated with the indicated SARS-CoV-2 variants. CPE formation was analyzed after 72 h. Average values of 16 biological replicates (except n=8 for bamlanivimab) are depicted. Using A549-AT cells, CPE formation was microscopically evaluated. Physiological concentrations of mAbs were applied according to manufacturer’s instructions.

In agreement with previous studies both Epsilon variants were less susceptible towards imdevimab, but were efficiently neutralized by casirivimab alone or in combination with imdevimab. Of note, using imdevimab or the combination of casirivimab/imdevimab, the B.1.429 strain used in this study was less susceptible when compared to B.1.427 (Fig. 2). In conclusion, all SARS-CoV-2 variants carrying L452R were resistant to bamlanivimab. Even at high concentrations imdevimab was not effective against Delta indicating high resistance, but only a moderate reduction in neutralization of all L452R variants was observed for the treatment with the clinically approved combination of casirivimab/imdevimab.

## 4. Discussion

The emergence of SARS-CoV-2 variants of concern which in addition to increased transmission rates might be less sensitive to neutralizing antibodies challenge the health systems worldwide. The ongoing evolution of SARS-CoV-2 requires constant characterization of emerging mutations with regard to the efficacy of neutralizing mAbs. For the Alpha variant, which became dominant in late 2020, it could be shown that the N501Y substitution in S is associated with higher affinity for the ACE2 receptor and higher transmissibility [2, 25-27]. Nevertheless, the neutralizing ability of Alpha by convalescent sera or BNT162b2 was preserved towards authentic viruses [10, 28]. The global displacement of Alpha and the dominance of Delta since spring 2021 shifted the focus towards L452R-carrying variants Epsilon and in particular the lineage B.1.617. In this study, we observed a reduced sensitivity of variants carrying L452R towards convalescent and vaccine-elicited sera that was further diminished by substitutions at E484 and T478K in S, respectively (**Fig. 1**). Hence, the B.1.617 lineage (Kappa and Delta) exhibited a stronger immune escape relative to Epsilon.

The position E484 on the receptor binding domain (RBD) in S is a well-described immunodominant site [6, 29-31]. In this work, we detected a lower capacity of convalescent or vaccine-elicited sera to neutralize the E484Q-carrying variant Kappa matching the previously shown reduced neutralization of variants Beta and Zeta carrying E484K (Widera et al. 2021). Hence, both substitutions E484K and E484Q might limit the sensitivity of SARS-CoV-2 to neutralizing antibodies.

Specifically, using convalescent but not vaccine-elicited sera Kappa was neutralized 1.5-fold less effective than the T478K-carrying Delta variant. The T478K substitution appears more relevant for neutralization by convalescent sera. Thus, or data suggests that mRNA-based vaccination confers better protection against both B.1.617 strains when compared to convalescent sera.

The capacity to neutralize the L452R carrying Epsilon lineages was only moderately reduced for casirivimab and imdevimab (**Fig. 2**). Using pseudoviruses the L452R substitution was recently described as sensitive to both mAbs casirivimab and imdevimab, but has also been associated with increased viral shedding *in vivo* and reduced neutralization of RBD-and N-terminal Domain (NTD) antibodies [13].

Our data indicates that in contrast to casirivimab, imdevimab is less affected by substitutions at position E484 and thus effectively neutralizes Kappa and Epsilon. Based on structural comparisons of human SARS-CoV-2 neutralizing antibodies, casirivimab belongs to class 1 antibodies blocking ACE2 and binding to exclusively the ‘up’ RBDs of S [32]. Imdevimab, classified as class 3 antibody, binds outside the ACE2-binding site matching the here observed high sensitivity to Kappa and efficient neutralizing of E484K carrying variants Beta and Gamma, as documented recently [14, 33]. Hence, the combination of casirivimab and imdevimab confers unrestricted protection in variants carrying substitutions at L452 and E484.

In discrepancy to our study a recently published work by Planas and collegues [10], showed no reduction in the neutralization capacity of casirivimab or imdevimab for Delta relative to B.1. Our assays, however, revealed a substantial loss in the neutralization capacity of imdevimab against Delta. This was in agreement with another recent study demonstrating a >10-fold resistance of Delta towards imdevimab and further receptor binding motive (RBM) binding antibodies suggesting that both the L452R and T478K substitutions reduce the neutralizing activity [34].

The reduced imdevimab susceptibility towards Delta compared to Kappa indicates that T478K possibly in combination with other mutations found in Delta S might mediate the observed immune escape. Although imdevimab was predicted to bind S distal to the ACE2-binding site and potentially hinders receptor binding via steric interference [32]. Most likely, T478K could affect imdevimab neutralizing capacity, however, more studies are needed to evaluate the impact of T478K on S secondary structure and antibody binding. Both discussed studies [10, 34] were conducted with authentic viruses and are methodically comparable to our work. Since Delta exhibits a partial resistance towards imdevimab, the resistance barrier to the combination therapy of casirivimab and imdevimab together is also reduced. Consequently, a further single mutation in S of Delta could hamper the clinical efficacy of mAb combination therapy of casirivimab and imdevimab.

## 5. Conclusions

In contrast to the parallel circulating Kappa variant, the neutralization efficiency of convalescent and vaccine-elicited sera against Delta was only moderately reduced. SARS-CoV-2 Kappa variant harboring E484Q mediate resistance to casirivimab while the substitution T478K present in Delta results in limited neutralization by imdevimab. However, a combination therapy with imdevimab and casirivimab together is still effective against both B1.617 variants.

## Data Availability

All sequences are available on GenBank under accession numbers MT358638, MT358641, MT358643, MZ427280, MZ317895, MZ317896.2, MZ315140, and MZ315141.

## Funding

This study has been performed with support of the Goethe-Corona-Fund of the Goethe University Frankfurt (MW) and the Federal Ministry of Education and Research (COVIDready; grant 02WRS1621C (MW).

## Institutional Review Board Statement

The study was conducted according to the guidelines of the Declaration of Helsinki, and approved by the Institutional Review Board of the Ethics Committee of the Faculty of Medicine at Goethe University Frankfurt (250719).

## Acknowledgments

We are thankful for the numerous donations to the Goethe-Corona-Fund and for the support of our SARS-CoV-2 research. The authors would also like to thank all technical staff involved in data acquisition.

## Conflicts of Interest

SC was member of a clinical advisory board for Biontech. TW received speaker and consultancy fees from Gilead Sciencens, Merck Sharp Dome and Janssen Pharmaceuticals. All other authors declare no conflict of interest.

## Notes

### Funding Statement

This study has been performed with support of the Goethe-Corona-Fund of the Goethe Univer-sity Frankfurt (MW) and the Federal Ministry of Education and Research (COVIDready; grant 02WRS1621C (MW).

### Summary of Updates

Figure 1 and corresponding legend revised to clarify the basis of calculation for the indicated x-fold changes; statistical analysis updated for figure 1a; minor correction of typing errors; author affiliations updated; page numbers added.

